# History of premorbid depression is a risk factor for COVID-related mortality: Analysis of 1,387 COVID+ patients

**DOI:** 10.1101/2020.12.17.20248362

**Authors:** Sean A. P. Clouston, Benjamin J. Luft, Edward Sun

**Author notes:** **Correspondence to:**. Physical address: Sean Clouston, Program in Public Health, 101 Nichols Rd., Stony Brook Health Sciences Center, Stony Brook University, Stony Brook, NY, 11794-8338.

## Abstract

**Background:** The goal of the present work was to examine risk factors for mortality in a 1,387 COVID+ patients admitted to a hospital in Suffolk County, NY.

**Methods:** Data were collated by the hospital epidemiological service for patients admitted from 3/7/2020-9/1/2020. Time until final discharge or death was the outcome. Cox proportional hazards models were used to estimate time until death among admitted patients.

**Findings:** In total, 99.06% of cases had resolved leading to 1,179 discharges and 211 deaths. Length of stay was significantly longer in those who died as compared to those who did not p=0.007). Of patients who had been discharged (n=1,179), 54 were readmitted and 9 subsequently died. Multivariable-adjusted Cox proportional hazards regression revealed that in addition to older age, male sex, and heart failure that a history of premorbid depression was a risk factors for COVI-19 mortality (HR = 2.64 [1.54-4.54] P<0.001), and that this association remained after adjusting for age and for neuropsychiatric conditions as well as medical comorbidities including cardiovascular disease and pulmonary conditions. Sex-stratified analyses revealed that associations between mortality and depression was strongest in males (aHR = 4.45 [2.04-9.72], P<0.001), and that the association between heart failure and mortality was strongest in participants aged <65 years old (aHR = 30.50 [9.17-101.48], P<0.001).

**Interpretation:** While an increasing number of studies have identified a number of comorbid medical conditions and age of patient as risk factors for mortality in COVID+ patients, this study reports that history of depression is a risk factor for COVID mortality.

**Funding:** No funding was received for this study.

---

The SARS-CoV-2 pandemic has caused a global emergency causing millions of infections and cases of SARS-CoV-2-related disease (COVID-19) with New York City being an early epicenter in the U.S. (1). SARS-CoV-2 is a highly infectious disease with reports of the mean reproductive rate to be 1.94 (1.83-2.06) and mean time until death of 20 days (17-24) since onset of symptoms (2). Suffolk County, a large suburban commuter community on Long Island where essential workers for New York City often reside, was hit early with large numbers of cases and deaths attributed to COVID-19 (3).

There is a rapidly growing body of research about the details of the disease’s structure and dynamics (4). To date, however, relatively few studies have identified pre-existing conditions that are associated with mortality among individuals who are COVID+. Known risk factors include aging (5) as well as a range of aging-related inflammatory conditions (6). However, the majority of pre-existing conditions do not survive age adjustment (7) leaving little other than indicators of disease severity among infected patients (e.g., cardiac injury (8) and hypoxia (9), or large vessel strokes in younger patients (10)) to determine severity of COVID-19.

There is increasing attention to the impact of COVID-19 and functional changes to the nervous system (11) as indicated by changes to neuropsychiatric symptomatology (12). Notably, infection with COVID-19 has now been linked with increased depressive symptoms (13), anosmia (14), cerebrovascular neuropathology (15), as well as peripheral immunologic changes and increased risk of stroke (16). Noting that research prior to the COVID-19 epidemic suggests that patients with elevated depressive symptoms have significant neuro- and systemic immune dysfunction and activation (17), an alternative hypothesis is that COVID-19 may be more severe in patients with depression.

While studies have argued that COVID-19 may cause distress both to those who are infected and those who are actively avoiding infection, no studies to our knowledge have identified premorbid psychiatric risk factors for COVID-19 mortality. The objective of the current study was to determine whether premorbid psychiatric conditions including, in particular, depression, were premorbid risk factors for mortality in an inpatient sample admitted for COVID-19 treatment. We hypothesized that depression would be associated with increased risk of COVID-related mortality after adjusting for age and sex and other confounders including chronic heart failure.

## Methods

Stony Brook University Hospital (SBUH) is the largest medical center on Long Island, NY and manages the only Level I Adult Trauma Center. SBUH began monitoring its first confirmed COVID-19 in early March. The current study examined outcomes among COVID+ patients admitted from 3/7-5/15/2020. Outcomes were censored at death or on 9/1/2020 to allow time for cases to resolve. Patients who died in the emergency room prior to admission or within one day of admission were not included in multivariable analyses.

Patients who died prior to admission ranged in age from 46-101 years, none were depressed, six were female, and had on average 1.75 comorbidities including COPD and atrial fibrillation but not depression or chronic heart failure. In cases of readmissions for patients who had been discharged, the date of first admission was retained and, if applicable, the new date of discharge or death was recorded. Age-stratified subgroup analyses were completed, and sensitivity analyses were completed examining risk factors only for patients whose treatment was completed.

### Measures

The outcome was days until death due among inpatients admitted for COVID-19 care. Pre-existing conditions were recorded at intake by emergency medicine clinicians. While intake forms recorded all previously diagnosed conditions, only comorbidities that occurred frequently enough to identify statistically significant relationships were examined. Specifically, we recorded pre-existing diagnoses of 1) neurological or psychiatric disorders including generalized anxiety, major depression, mental disability, and dementia; 2) cardiovascular conditions including atrial fibrillation, cardio-arterial disease, chronic heart failure, hyperlipidemia, hypertension, and stroke; 3) digestive diseases including obesity, diabetes mellitus, and gastroesophageal reflux disease; 4) pulmonary conditions including asthma, and chronic obstructive pulmonary disease; and 5) other diseases including chronic kidney disease and all-cause cancer. Demographic variables included age in years, and sex. Age was entered in years, rather than by 5-year age groups, because age categorization usually results in loss of information and attribution of age variance to aging-related conditions. We also identified patients with no premorbid conditions. Sensitivity analyses additionally considered accounting for the number of comorbid conditions, which in this population ranged from 0-19 (mean = 3.13 [2.82]) but since these analyses did not identify significant correlations, this variable was dropped from models presented.

While the goal of the present study was not to study the effects of in-hospital treatments, some have noted that intubated patients were at increased risk of mortality (18). Yet, mechanical ventilation was most likely to be used among individuals with the most severe disease in the early weeks of disease management. To examine the importance of adjusting for intubation, we also conducted multivariable analyses adjusting for whether an individual had been intubated during their inpatient stay.

The guidance regarding ventilator use to treat COVID-19 changed in April 2020. Since then, intubation rates have fallen drastically. Since mortality rates, and vulnerability to COVID-related mortality, may differ in intubated patients, we examined risk factors in intubated and non-intubated patients, and also compared risk factors in patients who had been admitted prior to April 15^th^ when ventilation was still being used regularly versus thereafter when ventilation was much less common.

### Ethics

The Internal Review Board at Stony Brook University approved this study (IRB#1586703). All study activities were performed in accordance with relevant guidelines and regulations.

### Statistical Analysis

Patient characteristics were reported using means and standard deviations or percentages (%). Student’s t-tests and χ^2^ tests were used to complete unadjusted analyses. Multivariable-adjusted analyses relied on Cox proportional survival modeling was used to estimate time until death among all admitted patients when adjusting for age (19); the exact method was used to account for ties (20). Power simulations determined that at least 1,300 patients were required to achieve multivariable-adjusted power estimates (α=0.05, power=0.80); *a priori* projections completed in April indicated May 15^th^ as study closing date. Follow-up was allowed for at least twice the expected mean length of hospital stay until at least 99% of cases had resolved. Proportional hazards assumptions were tested. Model concordance (C) was reported to determine predictive accuracy; C ranges from 0.50 (poor predictor) through 0.70-0.80 (very good accuracy) to 1.00 (perfect prediction). All analyses were completed using Stata 15.1/SE [StataCorp].

## Results

The current study examined outcomes among COVID+ patients (N=1,387; 211 deaths) admitted from 3/7-5/15/2020 (mean length of stay=13.00 [SD = 18.04] days). Outcomes were censored at death or on 9/1/2020 to allow time for patients to resolve. At time of analysis, 99.06% of cases had been discharged (n=1,179) or had died (n=211). Of remaining cases, half had been intubated and the mean length of stay was 94.13 [SD=30.59] days.

COVID+ patients had a range of premorbid physical conditions including atrial fibrillation, cardio-arterial disease, chronic obstructive pulmonary disease, heart failure, hyperlipidemia, hypertension, and stroke (**Table 1**). The mortality rate was 1.15% [1.01-1.32] per day. Age was a strong predictor of mortality in all models with each additional year of age associated with 4.55% [3.57-5.53] increase in the risk of mortality in these models. Notably, age alone was a very good predictor of the risk of mortality in this population (C = 0.74). The predictive power was similarly good in females (C=0.77) and males (C=0.72), and in both individuals with any comorbidities (C=0.72) and among those with without any comorbidities (C=0.84).

**Table 1.**
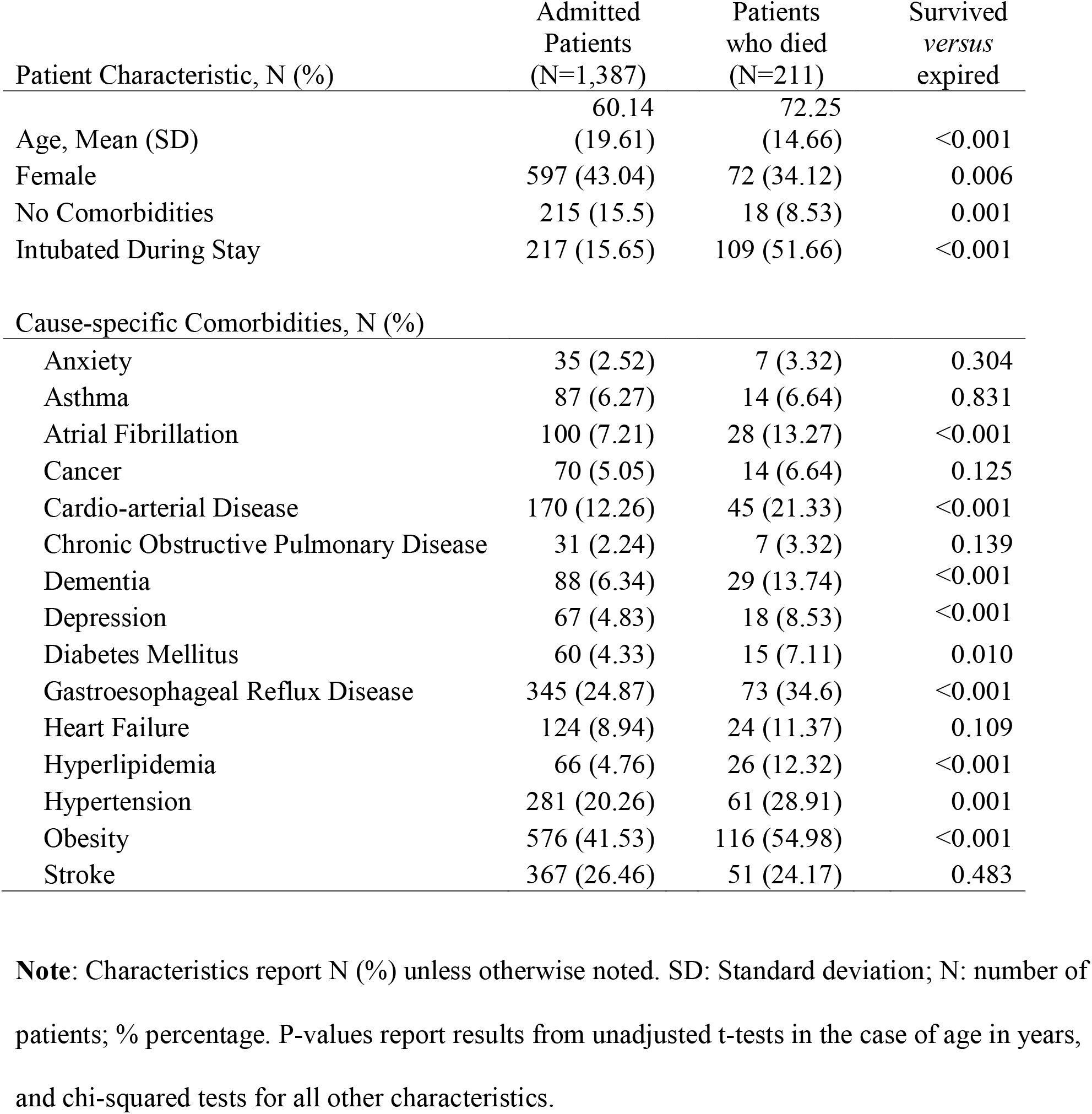
Patient characteristics for 1,387 patients admitted to Stony Brook University Hospital between March-May 15^th^, 2020

Length of stay was significantly longer in those who died (16.08 days) as compared to those who did not (12.43 days; difference=3.65 days, p=0.007). Of patients who had been discharged (n=1,179), 54 were readmitted (4.58% [3.46-5.93]) and nine (16.67% [7.92-29.29]) subsequently died. Seven patients died prior to, or within one day of, first admission. While these were excluded from multivariable analyses, these patients were older (78.76 versus 60.09, p=0.033) than patients who were included in the study.

Early efforts relied on ventilator usage to treat patients and in these data resulting in large differences in treatment course with, for example, 23.30% (n=103) of all admitted patients placed on ventilation in March 2020 versus only 10.73 (n=98) patients in April and May were intubated. Despite being a risk factor for morality, mortality rates were not lower in patients admitted in March as compared to either April (HR = 1.22, p=0.204) or May (HR = 1.50, p=0.131).

Age/sex-adjusted models identified intubation, chronic obstructive pulmonary disease, depression, and stroke as possible risk factors for COVID-19 mortality (**Table 2**). Diabetes and anxiety showed trends potentially indicative of risk factors. Additionally, analyses suggested that lacking any comorbidities was not associated with reduced risk of COVID-19 mortality. Multivariable-adjusted models revealed that in addition to previously described physical conditions, a history of depression was a significant risk factor for COVID+ mortality.

**Table 2.** Age/sex-adjusted and multivariable-adjusted hazards ratios and 95% Confidence intervals derived from Cox proportional hazards regression models examining predictors of mortality in COVID+ patients, Stony Brook University Hospital

| Patient Characteristic | Age/Sex-Adjusted |  |  | Multivariable Adjusted |  |  |
| --- | --- | --- | --- | --- | --- | --- |
|  | HR | 95% C.I. | P | aHR | 95% C.I. | P |
| No Comorbidities | 0.53 | (0.33-0.86) | 0.010 | 0.80 | (0.49-1.32) | 0.385 |
| Intubated | 1.91 | (1.43-2.57) | <0.001 | 2.33 | (1.72-3.17) | <0.001 |
| Pre-existing Medical Comorbidities |  |  |  |  |  |  |
| Anxiety | 1.79 | (0.83-3.84) | 0.136 | 1.59 | (0.74-3.43) | 0.237 |
| Asthma | 0.87 | (0.50-1.50) | 0.612 | 0.90 | (0.52-1.57) | 0.720 |
| Atrial Fibrillation | 2.24 | (1.5-3.36) | <0.001 | 1.23 | (0.81-1.86) | 0.339 |
| Cancer | 1.43 | (0.81-2.53) | 0.213 | 0.96 | (0.54-1.70) | 0.875 |
| Cardio-arterial Disease | 1.76 | (1.26-2.46) | 0.001 | 1.01 | (0.71-1.44) | 0.945 |
| Chronic Kidney Disease | 1.48 | (0.65-3.37) | 0.348 | 1.18 | (0.51-2.7) | 0.703 |
| Chronic Obstructive Pulmonary Disease | 2.61 | (1.75-3.89) | <0.001 | 1.77 | (1.18-2.65) | 0.006 |
| Dementia | 2.16 | (1.32-3.53) | 0.002 | 1.03 | (0.62-1.71) | 0.918 |
| Depression | 2.64 | (1.54-4.54) | <0.001 | 2.04 | (1.18-3.52) | 0.011 |
| Diabetes Mellitus | 1.49 | (1.12-1.99) | 0.007 | 1.38 | (1.03-1.84) | 0.032 |
| Gastroesophageal Reflux Disease | 1.69 | (1.09-2.62) | 0.020 | 1.28 | (0.82-2.00) | 0.277 |
| Heart Failure | 3.91 | (2.54-1.54) | 0.010 | 2.36 | (1.51-1.56) | <0.001 |
| Hyperlipidemia | 1.52 | (1.13-2.06) | 0.007 | 1.04 | (0.76-1.42) | 0.811 |
| Hypertension | 1.69 | (1.28-2.23) | <0.001 | 1.06 | (0.79-1.42) | 0.705 |
| Mental Disability | 2.37 | (0.58-9.76) | 0.232 | 2.24 | (0.54-9.28) | 0.266 |
| Obesity | 0.92 | (0.67-1.26) | 0.597 | 1.12 | (0.81-1.55) | 0.509 |
| Stroke | 2.45 | (1.55-3.87) | <0.001 | 1.35 | (0.84-2.17) | 0.217 |

**Note:** HR: Age/sex-adjusted hazards ratios; aHR: multivariable adjusted hazards ratio; 95% C.I.: 95% Confidence Interval. The proportional hazards assumptions were met in all models.

Stratified subgroup analyses (**Table 3**) in older (≤65 years) *versus* younger patients (<65 years) were substantively similar showing, for example, that depression was a risk factors for mortality in older patients though due to small numbers showed only a trend in younger patients. Subgroup analyses suggested that the association between depression and mortality was stronger in males than females. Sensitivity analyses did not identify factors that differed between intubated and non-intubated patients, and risk factors did not differ in the era of ventilation as compared to post-ventilation (after April 15^th^). Additionally, models considering the number of comorbid conditions did not change interpretation as compared to the model presented in Tables 3 or 4.

**Table 3.**
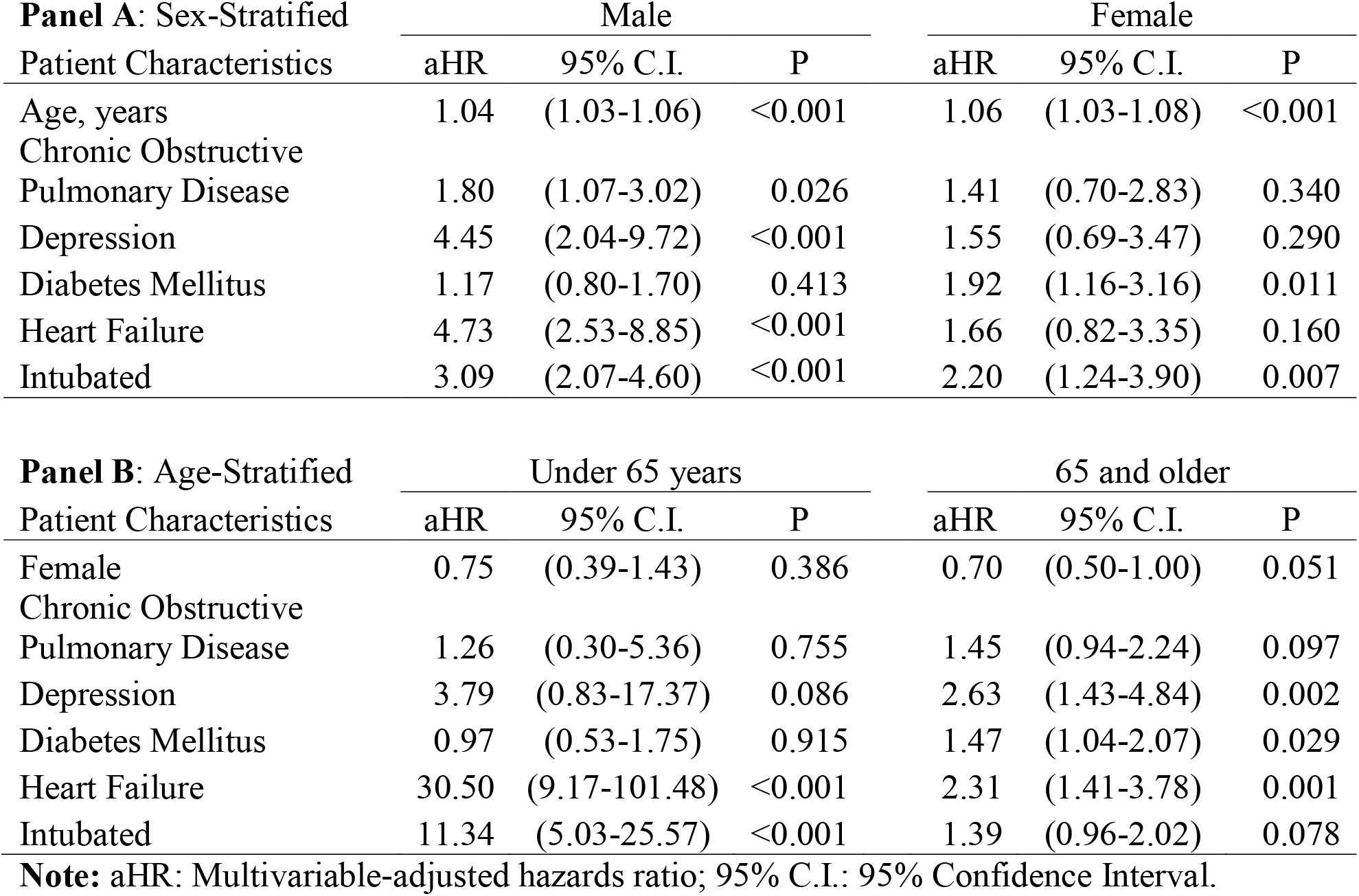
Multivariable-adjusted hazards ratios and 95% Confidence intervals derived from Cox proportional hazards regression models stratified by sex (Panel A) and by age (Panel B) examining predictors of mortality in COVID+ patients, Stony Brook University Hospital

## Discussion

This study sought to examine correlates of COVID-19 mortality. In the present study, we relied on a large cohort of patients admitted for COVID-19 in New York to determine that in addition to older age and male sex, a history of premorbid depression and heart failure were risk factors for COVID-19 mortality. Additionally, we determined that depression was a risk factor for COVID mortality in older patients, but that depression was most predictive in men as compared to women.

COVID-19 causes older infected individuals to experience more severe disease and large increases in risk of mortality (5, 21). Vulnerability to COVID-19 is a topic of intense interest that is important to individuals who may have an array of premorbid conditions and who may be making efforts to avoid infection. Prior work has highlighted a relatively large array of aging-related conditions such as diabetes and hypertension as potential risk factors for COVID-19. The current study supports earlier work suggesting that age is a substantively important risk factor for COVID-related mortality.

Individuals are increasingly being asked to return to work or else to justify staying home with many workers relying on comorbid conditions as a reason to justify working from home. The current study suggested that not all comorbidities confer increased vulnerability to COVID-19 mortality. However, we also found that lacking any comorbidities was not associated with reduced hazards of mortality among COVID+ patients. The present data suggest that while a number of comorbid conditions were associated with increased risk of COVID-related mortality in a medium-sized hospital in a large suburban community, that more research is warranted to better understand the mechanisms linking these risk factors to increased risk of mortality in these patients.

Our finding that depression is a risk factor for mortality in COVID+ patients supports prior analyses in a large population database (22). Mechanisms for such a relationship may include the potential for neuro- and systemic immune dysfunction that is common among individuals with depression (17). Indeed, patients with depressive symptoms have heightened levels of circulating proinflammatory cytokines including interleukin-6 (23), and heightened production of pro-inflammatory cytokines in response to stressors (24). This may be particularly important in the pathogenesis of COVID-19 given the purported crucial role that interleukin-6 plays in some initial reports of adverse outcomes related to COVID-19.

We did not find that hypertension was a risk factor for COVID-19 mortality, despite some reports to the contrary (25). Prior reports have suggested that there is some data to suggest that hypertension and diabetes both be risk factors because of their high prevalence in hospitals caring for COVID-19 (26). For example, in the largest such study (n=140), hypertension was described as being the most common comorbidity with a prevalence of 30% (27). Hypertension and diabetes are both high-prevalence age-related conditions, making them highly likely to present in a population of older adults. For example, the Centers for Disease Control and Prevention (28) estimate the prevalence of diabetes and hypertension in New York State to be 11.0% and 29.4% respectively, with estimates increasing to 24.1% and 55.6% respectively in respondents aged 65 and older. In the currently study (mean age 59.78), COVID+ patients had diabetes and hypertension at prevalence rates in this cohort that closely matched prevalence in the population, potentially highlighting the need for age-adjustment in research on studies examining predictors of COVID-19 severity and mortality.

Heart failure is a complex aging-related condition characterized by reduced functional capacity that carries reduced quality of life, increased healthcare usage, and high risk of mortality (29). Individuals with heart failure are at increased risk of dying from a range of conditions including from stroke and anemia, as well as from lung, liver, and kidney diseases (30). Heart failure is a common comorbidity in atrial fibrillation, a condition that often causes heart failure and also carries high risk of stroke (31). Together, these analyses support the increasingly common view that COVID-19 may cause a clotting disorder that accelerates mortality due to cardiovascular and cerebrovascular causes.

### Limitations

Though being among the earliest studies to identify pre-existing conditions that increase risk of mortality in COVID+ patients, this study is also limited in focusing on patients admitted to a single hospital on Long Island, NY. Information about race/ethnicity and socioeconomic status were not recorded. A number of studies have noted neuropsychiatric changes among COVID+ patients (12). NY. The current study did not seek to determine indicators of cardiovascular, pulmonary, or cerebrovascular indicators of COVID-19 severity but instead sought to identify patients for whom exposure to SARS-CoV-2 may be more deadly. Analyses did not identify dementia as a risk factor for mortality despite the fact that Alzheimer’s disease, a main cause of dementia, is a neuroinflammatory condition that causes elevated levels of c-reactive protein and cytokines commonly related to COVID-19. However, since individuals with Alzheimer’s disease and vascular dementia are most commonly cared for in nursing homes, where a large number of outpatient deaths are known to have occurred, this effect may be underestimated in studies such as this one that rely on inpatient samples. In the case that encephalopathy is present, but may not always become severe, future research is warranted to examine the potential for a lasting post-COVID encephalopathy among survivors. Major depression is a heterogeneous condition with a range of potential etiologies; we were unable here to examine whether subsyndromal depression or specific depressive symptoms were attributable for the increased risk identified herein. Due to limitations in the available data, we could not adjust for socioeconomic or racial/ethnic disadvantage here. Prior work suggests that these may play a significant role both in the risk of infection and in the risk of mortality. Further efforts are needed to characterize these risk factors for disease.

### Implications

There is increasing agreement that COVID-19 causes changes in the central nervous system (11) and that neuropsychiatric effects may be important when characterizing COVID-19 severity (32). There is also increasing recognition that these changes may carry long-term behavioral and functional consequences (33). The current study additionally suggests that those with pre-existing depression may also be at higher risk of experiencing the most severe forms of the disease.

## Data Availability

Data include private health information and are not available to outside users, but have been archived to COVID data-sharing websites including TriNetX.

## Declarations

### Ethics approval and consent to participate

The study was reviewed and approved by the Stony Brook Ethics Review Board; informed consent was not required because this was a secondary analysis of clinical data to help inform healthcare practices.

### Consent for publication

Not applicable.

### Availability of data and materials

All data include sensitive private health information. As such, data are being maintained by the PI but are also available to other researchers who are interested via the TriNetX data housing service.

### Methods

All methods were performed in accordance with the relevant guidelines and regulations.

### Competing interest

The authors have no conflicts of interest to disclose.

### Funding

No funding was received for this study.

### Author contributions

SC completed the analyses and drafted the paper. BL provided topical expertise and edited the manuscript. ES developed the hypotheses and provided scientific oversight.

## Acknowledgements

Not applicable.

## Notes

### Competing Interest Statement

The authors have declared no competing interest.

### Funding Statement

No funding was received for the completion of this study.

### Author Declarations

Stony Brook IRB Reviewed and approved the study (#1586703).

### Summary of Updates

Streamlining analysis.

